# Bone marrow from focal segmental glomerulosclerosis displays activation of inflammatory pathway

**DOI:** 10.1101/2023.03.06.23286859

**Authors:** Priyanka Rashmi, Patrick Boada, Arvind Soni, Tara K Sigdel, Dmitry Rychkov, Eunsil Hahm, Andrea Alice Da Silva, Izabella Damm, Rohan Paul, Flavio Vincenti, Jimmie Ye, Jochen Reiser, Jeffrey wolf, Minnie M. Sarwal

## Abstract

Circulating factors resulting from immune dysfunction have been proposed as one of the causes for increased risk of graft loss associated with recurrence of focal segmental glomerulosclerosis (FSGS) after kidney transplant. However, the precise identity of the circulating factors and their sources remain incompletely characterized. *In vivo* studies in mouse models have implicated a role for immature bone marrow cells in the development of FSGS. Using single-cell RNA sequencing we have profiled >50,000 cells from bone marrow of FSGS patients with or without recurrence after kidney transplant and controls including healthy individuals and patients with end-stage renal disease due to non-FSGS causes. Bone marrow mononuclear cells from patients with recurrence of FSGS after transplant display an inflammatory phenotype with activation of cytokine and interferon signaling in neutrophils, T cells and B cells. We also observe a dramatically depleted B cell population in R-FSGS patients. Conditioned media from BMNCs of R-FSGS patients have higher levels of pro-inflammatory cytokine MIP-1α/CCL3, reduced anti-inflammatory chemokine CCL22 and cause injury in a human podocyte cell culture model. Our studies provide evidence for the role of bone marrow cells in FSGS associated inflammatory milieu and elucidate the transcriptional changes associated with the disease.

## Introduction

Focal segmental glomerulosclerosis (FSGS) is a histopathologic lesion characterized by the scarring/sclerosis of some but on all glomeruli^1^. It accounts for approximately 20% of cases of nephrotic syndrome in children and 40% of those in adults^2^. The glomerular scarring is progressive and can be caused by a multitude of reasons including primary (idiopathic, no known associated cause), genetic (due to mutations in podocyte associated genes) and secondary (due to drugs, infections, or mal-adaptive alterations in kidney)^3, 4^. Primary FSGS is the most common cause of glomerular disease often leading to end stage renal disease requiring dialysis or transplantation. A major obstacle in kidney transplantation for primary FSGS is the recurrence of disease in the transplanted kidney increasing the risk of graft loss^5^. FSGS recurs in up to 60% of first kidney grafts and the risk of recurrence can be as high as 80% in patients who have lost their first graft due to recurrent FSGS (R-FSGS)^6^. A large body of clinical and experimental evidence including the fact that FSGS can recur in the transplanted kidney in hours to days after transplantation suggest that pathogenic factors in circulation in FSGS patients cause injury and recurrence of FSGS in the transplanted kidney are the pathogenic cause of recurrence^7-9^. Despite decades of studies, neither the identity of the circulating factor nor the source have been conclusively identified. Primary and recurrent forms of FSGS are most commonly treated with nonspecific therapies such as immunosuppressive drugs (glucocorticoids and calcineurin inhibitors), plasmapheresis or immunoadsorption^10^. An earlier study demonstrated that transfer of CD34^+^ hematopoietic stem/progenitor cells but not CD34^-^ peripheral blood mononuclear cells from nephrotic patients with glomerular disease caused proteinuria development and foot process effacement in a humanized healthy mouse^11^. This study suggested that the circulating factor is a product of immature differentiating cells. Later, another group confirmed this hypothesis in mice by demonstrating that the bone marrow myeloid lineage cells with low Gr-1 and Sca-1 expression are expanded during disease and leads to production of suPAR further aggravating the pathogenesis^12^. However, the equivalent cellular population in humans does not exist, still leaving the identity and source of the circulating factor associated with the pathogenesis of FSGS and R-FSGS in humans unknown. Here we have performed the transcriptomic profiling of FSGS patients with recurrence of disease after transplant and compared it to that of patients with no recurrence to characterize the undifferentiated cell population and transcripts altered in R-FSGS.

## Methods

### Bone marrow collection from patients and patient characteristics

BM aspirates from 8 FSGS patients and two control patients with end stage renal disease due to non FSGS related causes were collected with informed consent (IRB #17-22287). BM-aspirates of age and gender matched 5 healthy donors were purchased from Lonza Inc and used as healthy control. In the FSGS group, 5 patients had a transplant due to biopsy proven primary FSGS, followed by recurrence of FSGS within 3 months after transplant. Aspirate was collected while the patients are waiting for a new transplant. Aspirate was collected from 3 patients with biopsy proven primary FSGS at the time of first transplant and followed up for 6 months for the absence of recurrent FSGS (NR-FSGS). Among the control patients, one had ESRD associated with renal artery stenosis and one due to membranoproliferative glomerulonephritis (MPGN) and hypertension and bone marrows was aspirated at the time of transplant. Entire study cohort consisted of 7 male, 1 female FSGS patient; 3 male and 2 female healthy individuals and 2 male patients of ESRD due to non-FSGS causes. The mean age (years, mean ± stdev) of the R-FSGS, NR-FSGS and control cohorts are 38.8±6.6, 31.3±5.1 and 28.4±9.8 respectively with no significant differences among the groups.

### Isolation of bone marrow cells (BMC) and bone marrow mononuclear cells (BMNC)

One part of the bone marrow aspirate was centrifuged, and the pellet was treated with ACK lysis buffer to remove red blood cells and collect bone marrow cells (BMCs). BM mononuclear cells (BMNCs) were isolated by ficoll gradient centrifugation of the second part of aspirate and cryopreserved in 90% fetal bovine serum and 10% dimethyl sulfoxide (DMSO) at 5 million cells per vial.

### Single-Cell Preparation

On the day of experiment, cells were thawed, and single cell suspension was prepared in 2ul of phosphate-buffered saline (PBS) and 0.05% Ultrapure bovine serum albumin. Cell viability was assessed and cell populations with ≥80% viability score were used for the experiment. Cell number/volume was adjusted to the target before loading onto the 10X Chromium controller.

### Single Nucleotide Polymorphism (SNP) array

Genomic DNA was isolated from leftover cells or peripheral blood mononuclear cells of everyone in the cohort using QIAamp DNA Mini Kit (Cat# 51104, Qiagen, Hilden, Germany). We obtained ∼30ng of DNA from 5000 cells. DNA was used for genotyping using SNP arrays (OmniExpress Exome kit, Illumina).

### Library Preparation and Sequencing

Libraries for single-cell RNAseq were prepared using the 10X Single Cell Immune Profiling Solution Kit according to the standard manufacturer protocol.

Bone marrow mononuclear cells: For multiplexed RNAseq (Mux-Seq), 5,000 cells from 12 individuals of the cohort (3 R-FSGS, 3 NR-FSGS, 2 controls with ESRD due to non FSGS causes and 4 healthy individuals) were mixed to create a pool of 60,000 cells and loaded onto each lane. Three such pools were run. Additionally, BM-MNCs (50,000 cells) from two unique R-FSGS and one healthy individual were loaded onto each lane individually (Singleplex RNAseq). Full length cDNA libraries were prepared using the Chromium Single Cell 3′ Reagent Kits (v2): Single Cell 3′ Library & Gel Bead Kit v2 (PN-120237), Single Cell 3′ Chip Kit v2 (PN-120236) and i7 Multiplex Kit (PN-120262) (10x Genomics)^13^, and following the Single Cell 3′ Reagent Kits (v2) User Guide (manual part no. CG00052 Rev A). Libraries were run on an Illumina NovaSeq S2 as 150-bp paired-end reads. The minimum sequencing depth was 50,000 reads/cell using the read lengths 26bp Read1, 8bp i7 Index, 91bp Read2.

Data was processed via the 10X Chromium 3’ v2 platform. Data matrices and barcode information were generated using the 10X Cell ranger version 3.1.0 software, aligned to the GRCh38-3.0.0 transcriptome. After 10X generated unfiltered barcodes, the EmptyDrops tool was used to identify true cells with an FDR threshold of 0.01. In order to demultiplex each pool used for Mux-Seq and identify the original sample source for each cell, the computational tool-Freemuxlet was applied as described before (https://github.com/yelabucsf/popscle). Single nucleotide variants were annotated within the aligned reads from each pool using the 1000 Genomes variant database. A Bayesian clustering algorithm was applied to assign each cell to a group of cells with like genotype. Reference sample genotypes were determined using the Infinium OmniExpressExome-8 BeadChip (Illumina) and these were used to map each cluster to known subject identity. Freemuxlet is a genotype free adaptation of previously described tool Demuxlet^14^. Furthermore, Freemuxlet also detects and removes droplets containing two or more cells as determined by the presence of SNPs originating from more than one genotype^14^. Doublets and ambient cell readouts from singleplex dataset were removed from by using scrublet^15^ (0.2.3). Data from singleplex and multiplex runs were merged and a threshold for minimum expression of 200 genes per cell and percentage of transcriptome containing mitochondrial reads at 10% was set^16^. Genes were filtered for expression in at least 3 cells. MALAT1 was removed as a confounding marker prior to normalization^17, 18^. Experimental data was then normalized to a depth of 10,000, percent counts of mitochondria were regressed out, and the data was scaled. Batch effects between experiments were reduced by using ComBat^19^.

### Data Analysis

Data analysis was done using Python (3.8.8), Scanpy (1.8.2), R (4.2.0), and Seurat (4.1.1)^20-22^. Highly variable genes were identified for feature selection after data integration and normalization. A non-supervised Uniform Manifold Approximation and Projection (UMAP) was performed upon the dataset to cluster cells with similar transcriptional profiles. The Leiden clustering algorithm was used to cluster the distinct cellular neighborhoods at resolution 1.0^23^. Average expression of each gene was calculated for each cell cluster, then logged fold change was determined by taking the average expression of a single gene in one cluster against all other clusters. FDR-corrected p-values were calculated using the Wilcoxon rank-sum test. For cell type classification, we developed an automated prediction algorithm based on Human Cell Atlas bone marrow reference dataset^24^. The algorithm returns the identity of the cluster based on the frequency of genes associated with a cell type in the reference dataset and percentage of cells in the cluster expressing the respective genes. Predictions were manually curated for accuracy and authenticity based on previously validated markers. To understand the heterogeneity of a cell population, re-clustering was conducted by stratifying all neutrophils, T cells, B cells and CD34+ cells. Gene set enrichment analysis on differentially expressed genes with adjusted p value ≤ 0.05 was performed with gseapy (0.10.4)^25, 26^. Pseudotime computation and cellular trajectory was performed using the monocle2 library (2.24.0)^27, 28^. Data was made interoperable between python and R-with sceasy (0.0.6)^29^.

### In vitro culture of bone marrow mononuclear cells and assessment of secreted pathogenic factors

Mononuclear cells from patients and healthy controls were cultured to obtain conditioned media. Approximately, 300×10^3^ cells were plated in one well of 24 well plates and cultured in DMEM supplemented with 10%FBS and Penicillin/Streptomycin for 24h. After 24 hours 500ul of supernatant was collected and centrifuged to pellet the non-adherent cells. 300ul supernatant was removed, divided into 100ul aliquots and stored in -80°C until further use. Pellet was resuspended in remaining 200ul media and added back to the culture. After another 24h, supernatant was collected as before. Cytokines, chemokines and other inflammatory factors were quantified by previously described Meso Scale Diagnostic (MSD) based multiplex platform^30^. Immortalized human podocytes were cultured in six well plates and allowed to differentiate for 10-14 days. After differentiation, podocytes were either left untreated or treated for 24h with 200ul of BMC conditioned media collected after 24h and 48h of BMC culture. Podocytes were fixed in 4% PFA and 2% sucrose for 10min, permeabilized with 0.3% triton in PBS and then stained with rhodamine conjugated phalloidin to visualize stress fibers. Nuclei were stained with DAPI. Podocytes positive for stress fibers were quantified by manual counting after taking images at 40X using Leica SP5 confocal microscope.

## Results

### Single cell sequencing of bone marrow cells and mononuclear cells from healthy and ESRD patients

Mononuclear cells isolated from the bone marrow of 4 healthy individuals and 10 end-stage renal disease (ESRD) patients (5 recurrent, 3 non recurrent and 2 renal failures due to causes unrelated to FSGS, see study design **Figure 1**) were subjected to single cell transcriptomic profiling (10X genomics platform) as described in materials and methods. After quality control, we obtained 35,229 cells that included cells from all patients (**Figure S1**). Unsupervised clustering and projecting all cells in reduced dimension by uniform manifold approximation and projection (UMAP) to identify 26 clusters (**Figure 2A**). All clusters were annotated using previously established lineage markers and reference dataset^24^ in the unique gene expression profile of each cluster (**Table S1**). Therefore, we identified all major immune cell types including cells of myeloid, lymphoid and erythrocyte lineage (**Figure 2B**). We also identified two populations of CD34^+^ hematopoietic stem/progenitor cells (3,591 cells, 10.2% of all hematopoietic cells) making their proportion comparable to what is reported in the literature^24^. Percentages of the five major cell types between FSGS patients and controls were comparable except for B-cells which was significantly reduced in R-FSGS patients (**Figure 2E and 2F**).

**Figure 1.**
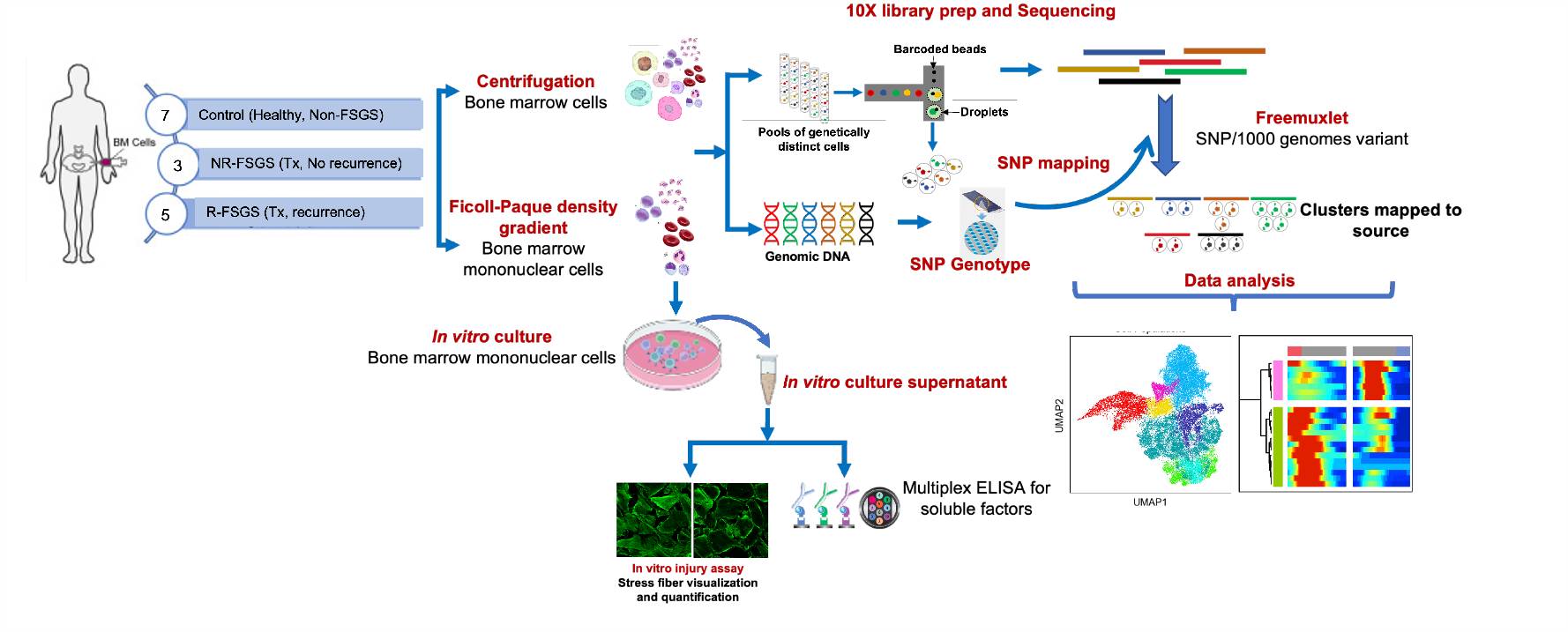
Study design. Bone marrow aspirate from patients with ESRD due to biopsy proven primary FSGS (n=10) or controls with ESRD due to other non-FSGS causes were collected at UCSF. Aspirates from 5 age and gender matched healthy individuals were purchased from Lonza. Aspirate was centrifuged to collect total bone marrow cells or enriched for mononuclear cells by Ficoll-Paque density gradient. Equal number of cells from each individual were pooled to load 60,000 cells per lane for cDNA library preparation on 10X platform using chromium single cell 3’ v2 chemistry. Cells were subjected to single cell RNA sequencing on Illumina NovaSeq S2 using 150bp paired end sequencing with minimum read depth of 50,000 reads per cell. Each individual in the cohort was genotyped using Illumina Omniexpress SNP array kit. Computational tool “Freemuxlet” with SNPs included from 1000 genomes variant was used to cluster the sequences followed by mapping to their original source using the genotypes. Data was analyzed in python to generate UMAPs and perform differential gene expression and trajectory analysis. Bone marrow mononuclear cells were cultured *in vitro* and supernatant was collected for the validation of secreted inflammatory factors by ELISA and podocyte injury assay.

**Figure 2.**
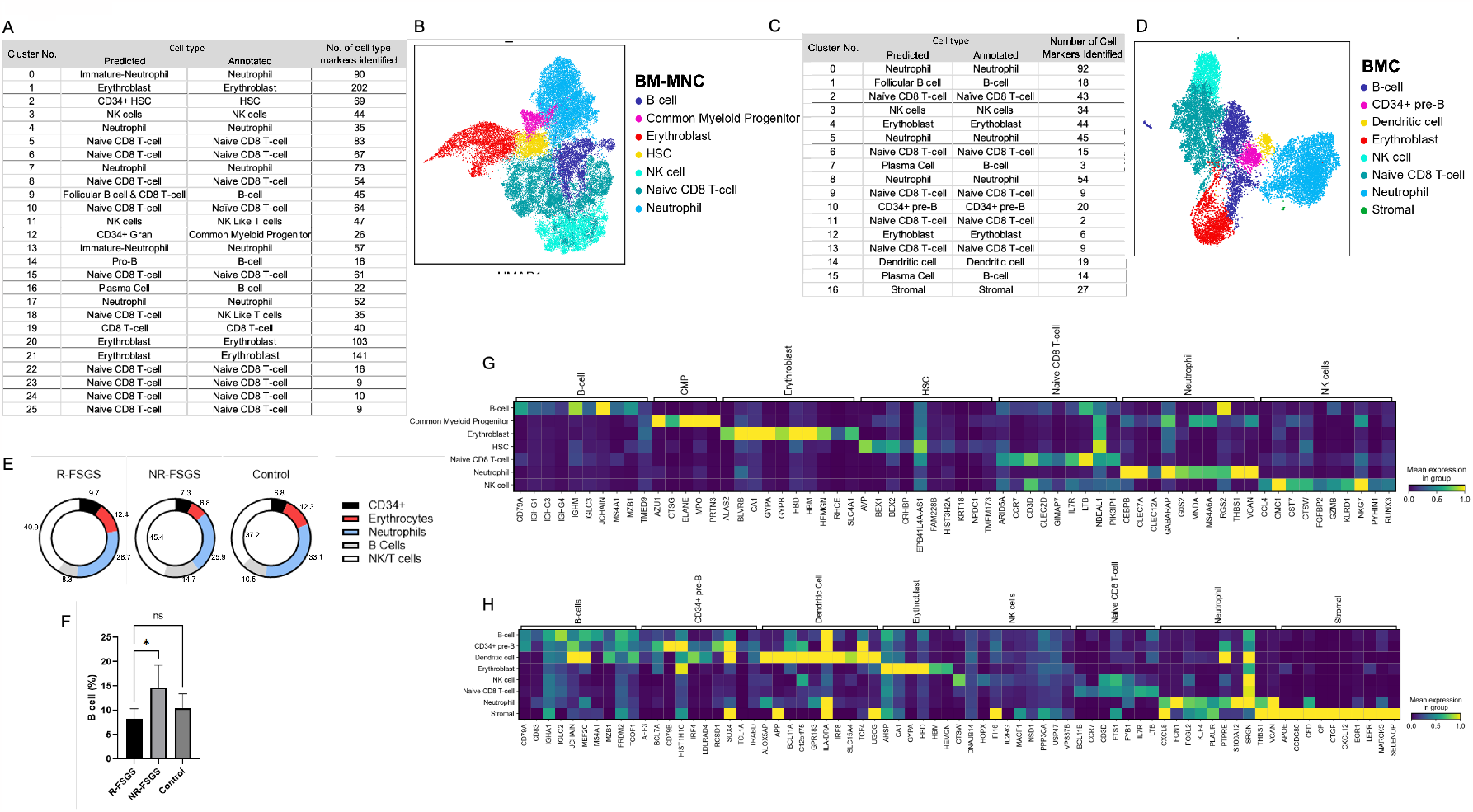
Single cell transcriptomes from bone marrow mononuclear cells and total bone marrow cells were analyzed by single cell RNA sequencing (scRNAseq). A) Unsupervised clustering of 35,229 mononuclear cells resulting in 26 distinct cell populations and annotated using canonical markers. B) UMAP displaying the distribution of 7 major cell types identified from (A). C) 15,376 single cell transcriptomes from all cells of bone marrow followed by unsupervised clustering resulted in 17 distinct cell populations. D) UMAP of 8 cell types including stromal cells identified based on canonical markers in (C). E) Proportions of individual cell populations in R-, NR- and control groups. F) B-cells are significantly reduced in R-FSGS. G and H) Top ten significantly expressed gene in each cluster of MNC and whole bone marrow cells has been shown to correlate with canonical markers of cell types assigned.

Additionally, to capture stromal cells of the bone marrow we centrifuged the aspirate and collected all cells for single cell RNA sequencing. This subset of patient cohort included 3 FSGS patients (2 R-FSGS and 1 NR-FSGS) and 5 controls consisting of 3 healthy individuals and 2 ESRD patients with renal failure due to non-FSGS causes. After initial processing and QC, we obtained 15,376 cells resolving into 17 clusters representing 8 cell types including one cluster of stromal cells (**Figure 2C, 2D**).

The top five genes from each cluster show their predominant expression in the cell type assigned to them and little to no expression in other cell types for bone marrow mononuclear (**Figure 2G**) as well as all cells in the bone marrow (**Figure 2H**). All clusters identified in the UMAP of BMNCs and BMCs consisted of cells from all patients suggesting minimal batch effect (**Figure S1**). UMAP of mononuclear cells from healthy individuals and patients with ESRD due to non-FSGS causes displays even distribution (**Figure S2A**) however, differential gene expression shows that there are overlapping genes between FSGS and non-FSGS patients (**Figure S2B**). There are 34 significantly upregulated genes in cells from non-FSGS patients compared to healthy individuals that included *CCL2, MAP3K8, CXCL2, CXCL8*, involved in inflammation via TNF and NF-κB signaling pathways. Two TNF pathway mediators tissue inhibitor of metalloproteinases-1 (TIMP-1) and monocyte chemoattractant protein-1 (MCP-1/CCL2) have recently been proposed to be a urinary biomarker in a cohort of FSGS patients^31^. Additionally, genes involved in hypoxia response such as *HMOX1, HIF1A*, and *IFNGR1* were upregulated in non-FSGS mononuclear cells (**Figure S2C**). These are well established non-specific response to injury and inflammation. Therefore, we used a combination of healthy and non-FSGS cells as control to identify unique transcriptional changes in R-FSGS patients.

### Transcriptomic signature of mononuclear cells in bone marrow of R-FSGS patients

Differential gene expression analysis was conducted to discover the identity of transcriptionally altered genes in bone marrow mononuclear cells of FSGS patients with respect to the control. The number of significant differentially expressed genes was 3,247 and 4,753 in R-FSGS and NR-FSGS MNCs respectively. 588 genes were unique to R-FSGS. Relative expression of several genes with established roles in the maintenance of renal structure and function were found to be altered. Among the top genes upregulated in R-FSGS patients were *ALDOA* (Aldolase, fructose-bisphosphate A), cytokines *CCL2* and *CCL7*, multiple HLA genes (HLA-A, B and C) involved in the antigen processing and presentation of endogenous peptides via MHC class I (**Figure 3A**). Interestingly, among the downregulated genes we observed CCL4 which is a major HIV-suppressive factor produced by CD8+ T-cells (**Figure 3B**). We have previously described a panel of autoantibodies in the pre-transplant sera of FSGS patients that can predict recurrence after kidney transplant with high accuracy with anti CD40 autoantibodies demonstrating the strongest correlation^30^. While we didn’t see any significant difference in CD40 transcript levels between R-FSGS and NR-FSGS patients, we observed a significant upregulation of genes downstream of CD40 signaling such as *NFKB1/2* and *TRAF3* (**Figure 3C**). Gene set enrichment with GO shows enrichment of biological pathways that represent activation of the inflammatory response mediated by cytokines such as interleukin 1 and 12, TNF and interferons. Additionally, signaling via three cell types were upregulated: antigen receptor mediated signaling, T cell receptor mediated signaling and neutrophil signaling (**Figure 3D and Table S2**). Several ribosomal proteins involved in protein targeting to ER were also upregulated and pathway analysis shows enrichment of signal recognition particle associated protein targeting to membrane pathway and p53 mediated apoptosis.

**Figure 3.**
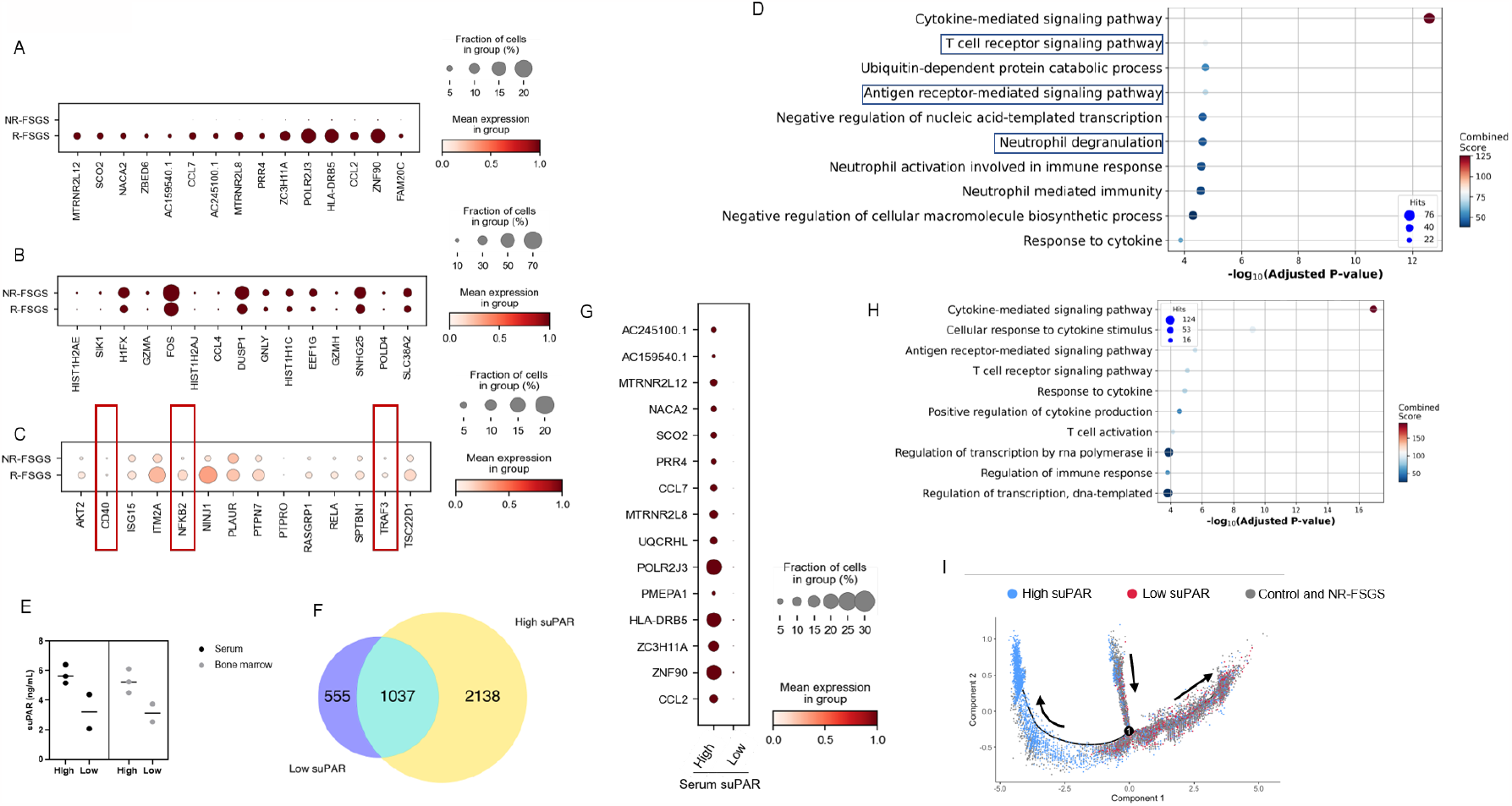
Transcriptomic differences between R-FSGS and NR-FSGS MNCs. A) Upregulated genes B) Downregulated genes C) Expression of FSGS associated genes D) Gene enrichment analysis using GO shows upregulation of pathways associated with B cells, T cells and neutrophils. E) Bone marrow aspirate and serum suPAR levels in R-FSGS patients separates the patients into two groups, with high and low suPAR respectively. F) Number of significantly altered transcripts in R-FSGS patients with high suPAR is approximately 4 times higher than that in patients with low suPAR. G) Dot plot representation of top ten significantly upregulated genes in R-FSGS patients with high suPAR. H) Gene enrichment of significant differentially upregulated genes in the high suPAR group revealed activation of B cell signaling I) Trajectory analysis of neutrophils showing separation of high and low suPAR cells at branch point 1.

### R-FSGS patients with high suPAR show upregulation of genes associated with B-cell activation and adaptive immunity

Prior studies have suggested that high suPAR in FSGS patients is associated with and causative of recurrence of FSGS^30^, however there is significant variability in serum suPAR levels in FSGS patients. Primary FSGS is most commonly treated with immunosuppressive drugs or plasmapheresis or immunoadsorption. Approximately 40-70% patients respond to therapy and this resolution is associated with reduction in serum suPAR levels^32, 33^. To understand if suPAR levels correlate with transcriptional changes in the bone marrow, we measured the serum suPAR in the R-FSGS patients and found that 3 patients have significantly high serum suPAR (**Figure 3E**) compared to the others. In order to understand the drivers associated with high suPAR, we compared the bone marrow transcriptional profile of R-FSGS patients with high serum suPAR levels to that of lower suPAR. Interestingly, we found that there were 2138 genes that were uniquely altered in bone marrow MNCs from R-FSGS patients with higher serum suPAR as opposed to 555 genes in R-FSGS patients with lower serum suPAR (**Figure 3F**). Significant upregulation of chemokines such as CCL2, CCL7, CXCL3, CXCL2 along with chemokine receptors CCR7 and CCL2 were observed (**Figure 3G**). Gene ontology analysis for functional enrichment revealed an upregulation of B cell receptor signaling and activation of adaptive immunity (**Figure 3H**). Neutrophils have been proposed as the primary source of shed suPAR during systemic inflammation^34^ and immature myeloid progenitors in mice were found to be sources of suPAR^12^. We looked at neutrophils and the transcriptional changes during neutrophil maturation using pesudotime analysis and found that the neutrophils from R-FSGS patients with high serum suPAR separate from the ones with low serum suPAR at branchpoint 1 of the trajectory along with the control cells (**Figure 3I and 4F**). The neutrophils from R-FSGS patients with high suPAR have elevated transcripts of genes associated with neutrophil activation and aggregation (**Figure 4G**). These results suggest that high suPAR levels in R-FSGS patients are associated with an elevated B-cell and neutrophil response.

**Figure 4.**
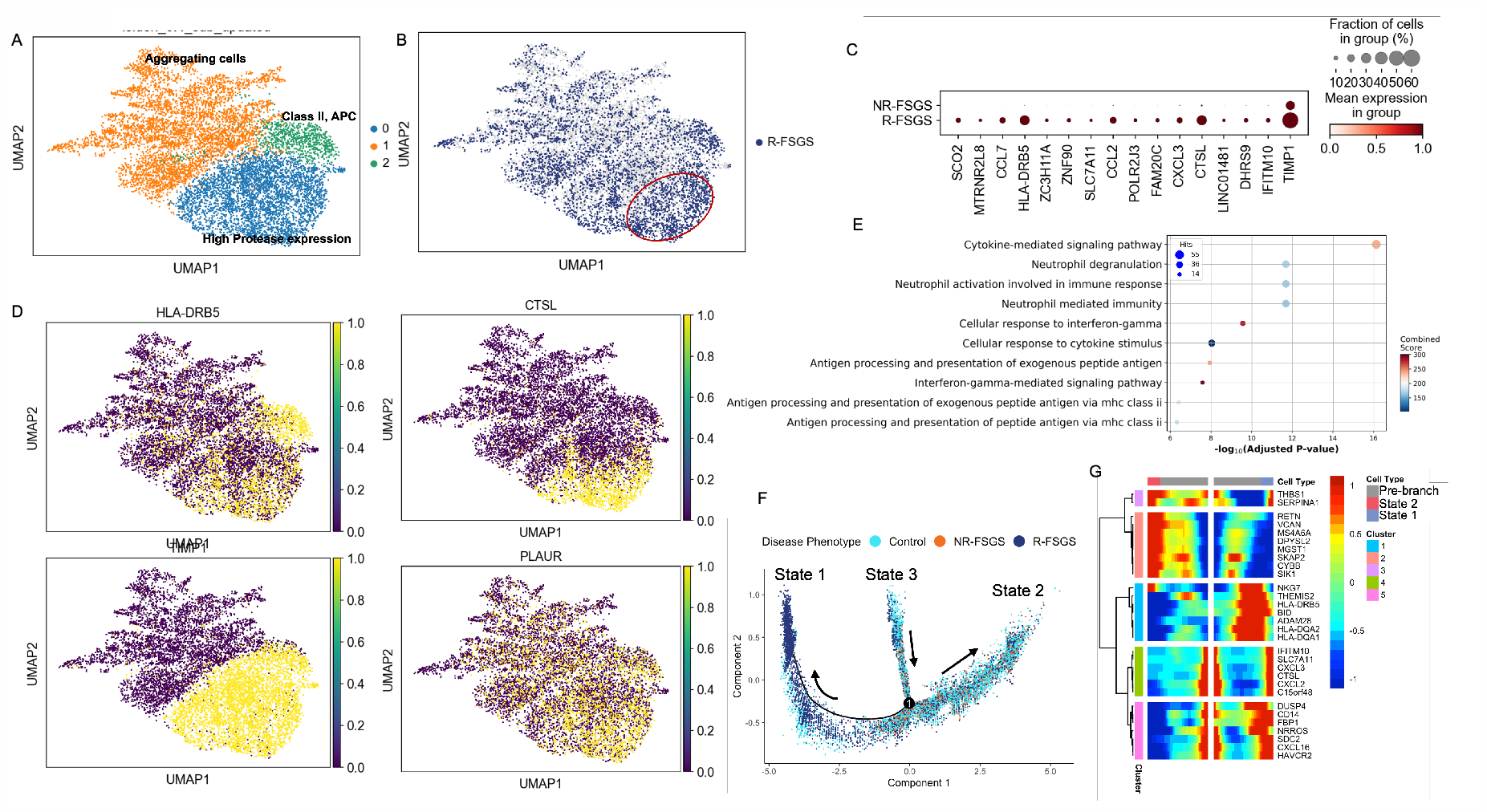
Unsupervised analysis of neutrophils shows downregulation of differentiation markers. A) Neutrophils in bone marrow resolve into 3 independent subclusters. B) Cluster 0 displays enrichment of bone marrow MNCs from R-FSGS patients. C) Genes with highest fold increase in neutrophils from R-FSGS patients. D) MNCs from R-FSGS patients in cluster 0 display upregulation of proteases CTSL, TIMP1 and MHC class II. E) Gene set enrichment with GO shows activation of neutrophils and inflammation associated phenotype. F) Pseudotime trajectory shows neutrophils from control and NR-FSGS patients (state 2) transcriptionally diverge from neutrophils from neutrophils from R-FSGS patients (state 1). G) Heatmap showing genes associated with the transcriptional changes between cells of state 1 versus state 2.

### Neutrophils show upregulation of proteases associated with podocyte injury

We did not observe any significant differences in the proportion of neutrophils of MNC from R-FSGS, NR-FSGS and control cells (25%, 26% and 30% respectively). Unsupervised clustering of neutrophils resulted in three populations (**Figure 4A**). the largest population (cluster 0) had high expression of proteases such as *CTSL* and *TIMP2* along with enrichment of cells from R-FSGS patients (**Figure 4A, 4B and 4D**). Cluster 1 population displayed activation of neutrophil aggregation and high expression of high expression of inflammatory markers such as S100 subunits (A8, A12, A9), VCAN and FOS (**Figure 4B and 4D**). Cluster 2 consists of a small population of APCs with high expression of class II MHC genes (**Figure 4A and 4D**). Top genes with significant upregulation in expression included chemokines *CCL7, CCL2* and *CXCL3* along with several proteases as mentioned above (**Figure 4C**). Gene set enrichment of significantly upregulated genes in neutrophils of R-FSGS patients demonstrated an activation of neutrophils and glycolysis (**Figure 4E**).

Pseudotime trajectory of neutrophils clearly revealed transcriptional changes resulting in the formation of two different cellular states. State 2 is enriched in control cells and neutrophils from NR-FSGS patients while state 1 is almost entirely composed of cells from R-FSGS patients (**Figure 4F**). Heatmap of significant genes associated with the transition at branch point 1 included activation of genes involved in LPS mediated inflammatory response such as interferon inducible *IFITM10* and *CXCL3* for the differentiation to state 1 (**Figure 4G**).

### The differentiation of T cells from R-FSGS patients is altered at transcriptional level

The T cells resolved into 6 different populations upon unsupervised analysis consisting of-naïve cells, T follicular helper cells, antigen presenting cells, cytotoxic T lymphocytes, cells with upregulation of SRP-associated protein translation and secretion, and a small cluster of apoptotic cells (**Figure 5A**). The cell population in cluster 2 was almost entirely composed of T cells from R-FSGS patients and the significantly expressed genes were enriched in antigen presentation of class I peptides (**Figure 5B**). We performed pesudotime trajectory analysis after combining T cells from all clusters. Genes specific to T-cell lineage differentiation were enriched in pathways associated with immune system process, leukocyte differentiation and activation among others (**Figure 5D**). The trajectory displays two distinct lineages of T cells that split at branch point 1 where T cell lineage from R-FSGS patients separate from that of NR-FSGS and control (**Figure 5E**). The two genes whose transcription level was dictating lineage included T-cell differentiation antigen CD6, and basic leucine zipper transcriptional factor ATF-like; AP-1 family transcription factor (BATF). CD6 is a cell adhesion molecule that mediates cell-cell contacts and regulates T-cell responses via its interaction with ALCAM/CD166. BATF controls the differentiation of lineage-specific cells in the immune system: specifically mediates the differentiation of T-helper 17 cells (Th17), follicular T-helper cells (TfH), CD8(+) dendritic cells and class-switch recombination (CSR) in B-cells. Other gene transcription responsible to dictate R-FSGS included, TRCB1 (T cell Receptor C Beta 1) which is cell surface marker in identification of T cell subsets, PERP (P53 apoptosis effector related to PMP-22), a protein with a role in stratified epithelial integrity and cell-cell adhesion, and AQP3 (Aquaporin-3), a water channel protein required to promote glycerol permeability and water transport across cell membranes.

**Figure 5.**
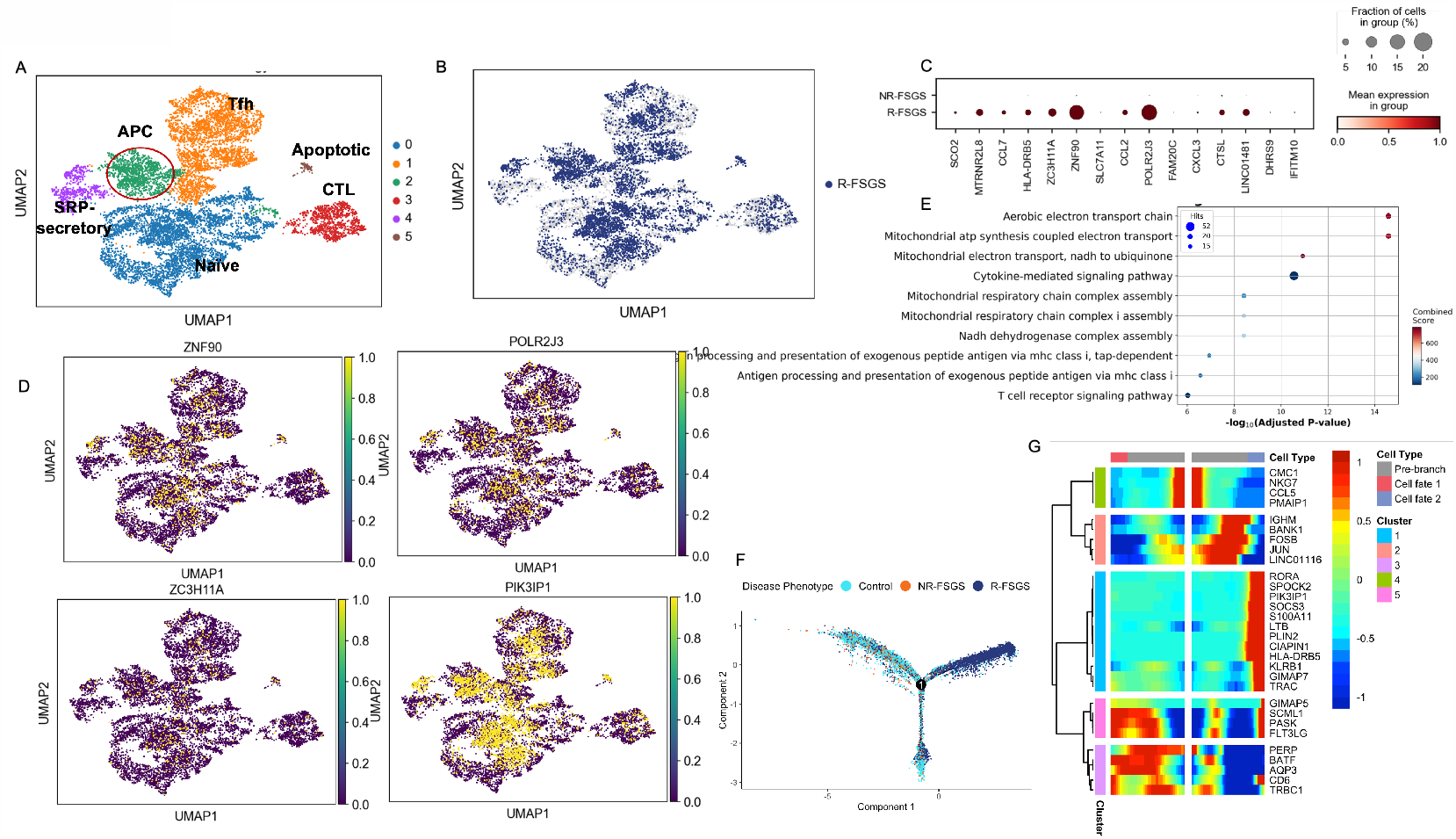
Unsupervised analysis of T cells. A) UMAP shows 6 different populations of bone marrow T cells. B) Cluster 2 and Cluster 5 show enrichment of T cells from R-FSGS patients. C) Gene enrichment of differentially expressed genes in T cells from R-FSGS vs NR-FSGS patients using GO platform. D) Pseudotime trajectory using monocle 2 shows T cells from R-FSGS patients separate at branchpoint 1 from the control cells based on their transcriptional profiles. E) Heatmap showing top genes associated with the developmental changes in transcripts from R-FSGS and branch containing cells from control and NR-FSGS T cells.

### B cells subclusters

Results described in Figure 3 and 4 both strongly suggest an involvement of B cells in the pathogenesis of recurrence of FSGS. Hence, we separated all B cells and performed an independent unsupervised analysis. Three populations of B cells resolved corresponding primarily to pre B, pro B and plasma cells (**Figure 6A**). While, we did not observe any enrichment of patient specific B cells in a particular cluster (**Figure 6B**), several genes were upregulated in B cells from R-FSGS patients including genes involved in transcription regulation such as *ZC3H11A, POLR2J3*; cytokines *INFNG2* (interferon gamma 2), *CCR7* and *SOCS3* (**Figure 6C**). Gene enrichment analysis using the subset of genes upregulated genes in B cells of R-FSGS patients showed enrichment of several pathways that included cytokine mediated signaling, negative regulation of apoptosis and antigen receptor mediated signaling (**Figure 6E**).

**Figure 6.**
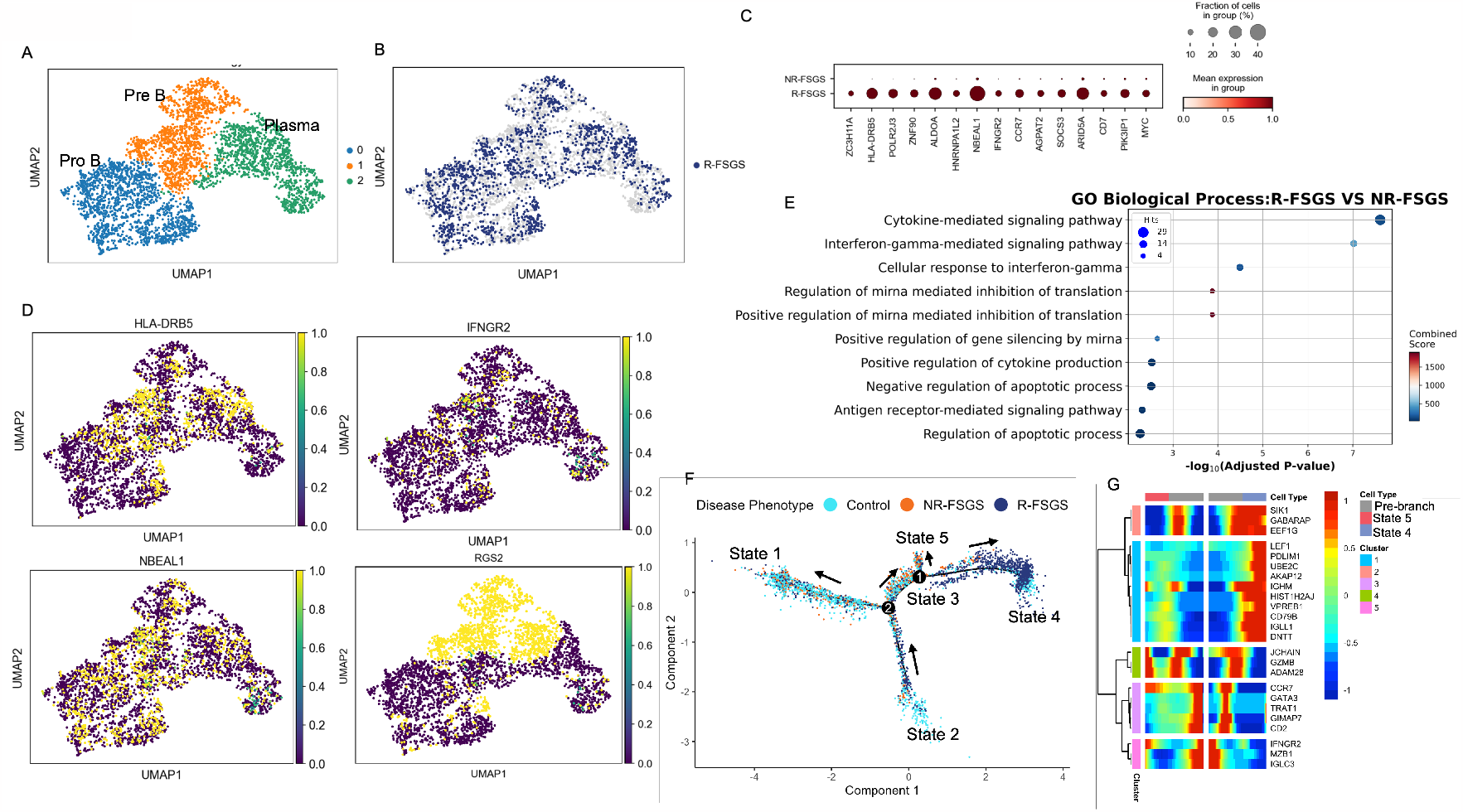
Unsupervised analysis of B cells in bone marrow. A) Three populations of B cells expressing markers of Pre B, Pro B and plasma cells were distinguished. B) B cells from R-FSGS were found throughout the bone marrow MNC landscape. C) Upregulated genes in B cells from R-FSGS patients compared to that in NR-FSGS. D) Expression on genes upregulated in B cells from R-FSGS superimposed on the UMAP. E) Gene enrichment analysis reveals enrichment of several pathways in B cells of R-FSGS patients including upregulation of cytokine signaling. F) Pesudotime trajectory displays transcriptional changes in B cells leading to the phenotype observed in R-FSGS patients enriched in state 4. G) Heatmap showing transcriptionally altered genes at branch point 1.

In order to further understand the altered trancriptonal profile of B cells in R-FSGS patients, we pooled the entire B cell population and performed a pseudotime analysis using Monocle 2. As seen in **Figure 6F**, B cells from all disease phenotypes are in state 2 assigned as the earliest pseudotime. At branchpoint 2, a major proportion of B cells from control and NR-FSGS patients separate and are in state 1 while rest of the cells transition primarily at branchpoint 1 resulting in state 4 cells which are almost entirely composed of B cells from R-FSGS patients. Heatmap in **Figure 6G** represents the predicted transcriptional regulators of transition from control/NR-FSGS B cells to the state enriched for B cells from R-FSGS patients. These cells show reduced expression of gene associated with B cell differentiation and maturation clustered in groups 3, 4 and 5. These include *J chain, MZB1* and *IGLC3* expression. Instead, B cells from R-FSGS patients show activation of genes associated with apoptosis induction and early stages of B cell differentiation associated with in the pre-B cell receptor complex (*VPREB1, CD79B, IGHM*).

### Differential gene expression profile of CD34+ cell population in R-FSGS vs NR-FSGS

Previous studies have shown that introduction of whole PBMC but not CD34 depleted PBMC from R-FSGS patients induces proteinuria in immunodeficient mice suggesting that the source of pathogenic factors in R-FSGS are the CD34+ cells^11, 12^. Therefore, to better understand the early progenitors in FSGS, we selected all cells in the CD34+ clusters (clusters 2 and 12) and performed an independent unsupervised analysis resulting in four individual sub clusters (**Figure 7A**) corresponding to hematopoietic stem cells (HSC), lymphocyte primed multipotent progenitors (LMPP), megakaryocyte erythrocyte progenitors (MEP) and granulocyte monocyte progenitors (GMP). Enrichment of bone marrow MNCs from R-FSGS patients was observed in cluster corresponding to HSC and LMPP (**Figure 7B**). Several genes were found to be upregulated in CD34+ cells from R-FSGS patients including genes involved in transcriptional regulation-*POLR2J3, ZNF90, ZC3H11A* (**Figure 7C**). Expression of one of the proposed pathogenic factors, *PLAUR* appears to be restricted to GMP cells (**Figure 7D**). Gene set enrichment of upregulated genes in CD34+ cells of R-FSGS patients with respect to NR-FSGS patients again shows enrichment of inflammatory signaling via cytokines and interferon gamma (**Figure 7E**).

**Figure 7.**
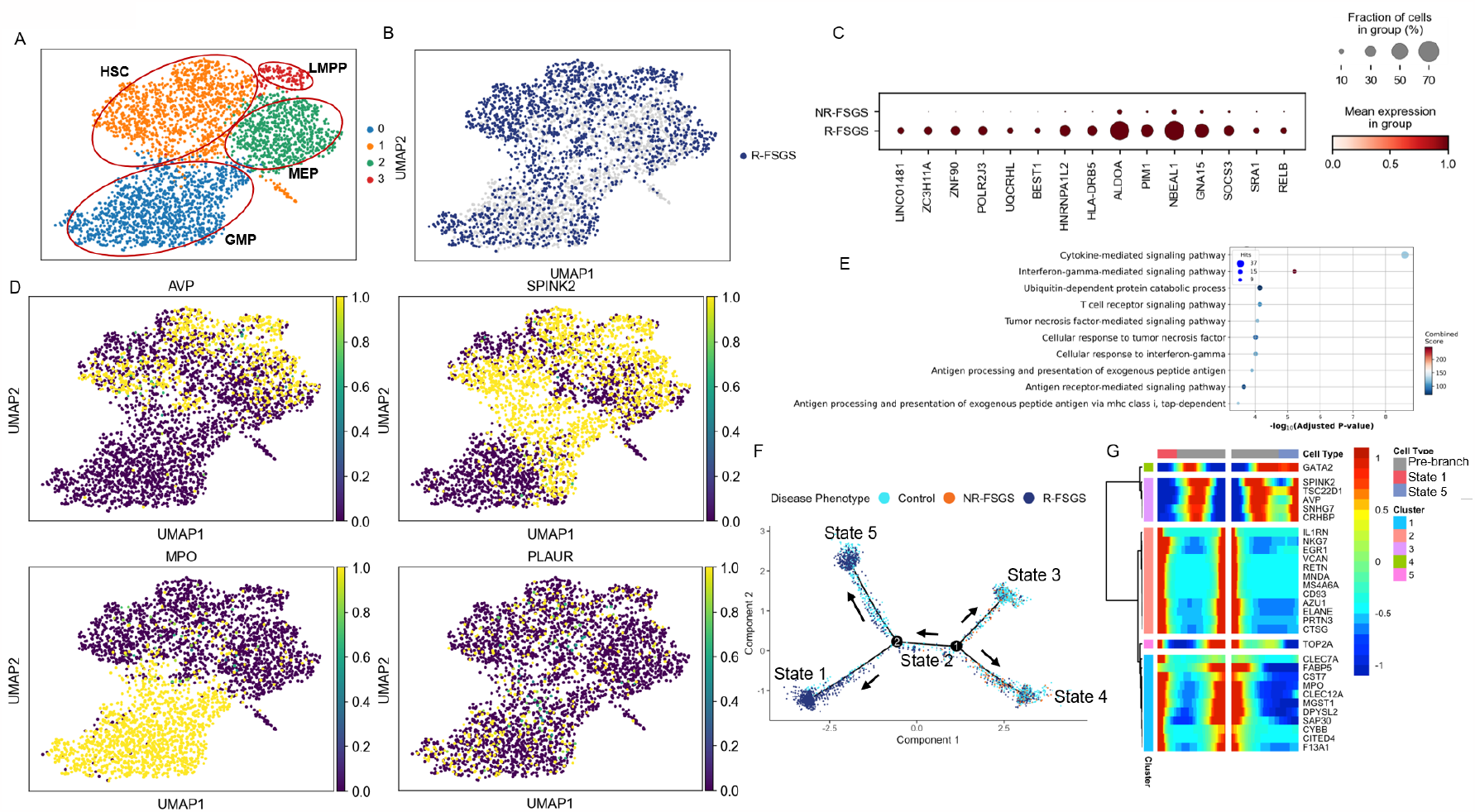
Unsupervised analysis of CD34+ cells. A) CD34+ cells in bone marrow resolve into 4 independent subclusters. B) Cluster 1 displays enrichment of bone marrow MNCs from R-FSGS patients. C) Dot plot showing top genes with significantly higher expression in CD34+ cells from R-FSGS patients. D) UMAP distribution of cells expressing several genes known to be important in progenitor cells. E) Functional enrichment of gene set obtained from differential gene analysis between R-FSGS and NR-FSGS cells and the pathways enriched. F) Trajectory obtained from pseudotime analysis shows control cells differentiating from cells from R-FSGS patients at branch point 2. G) Significant genes expressed before and after the branching at node 2 associated with the two different cell fates. LMPP: Lymphoid primed multipotent progenitor, MPP: hematopoietic stem cell/multipotent progenitor cell, MEP: megakaryocyte-erythrocyte progenitor cell, GMP: granulocyte monocyte progenitor cell.

Pseudotime trajectory analysis shows five different states of cells that transcriptionally diverge at two different branch points. At pseudotime 0, cells are in state 3 comprising of cells from all three groups. At branch point 1, cells separate into two states-state 4 containing all three groups and state 2 containing mostly cells from control and R-FSGS patients. At branch point 2, state 5 and state 1 separate where state 1 consists of R-FSGS and control cells in equal proportion but state 5 is almost exclusively CD34+ cells originating from R-FSGS patients.

### Culture of BMNC and measurement of secreted inflammatory mediators

As stated earlier, the etiology of FSGS can be diverse and in many cases, unknown called primary FSGS. However, the initial injury is primarily directed at podocytes^35^. To test if bone marrow in R-FSGS patients produces factors that can cause direct podocyte injury, we cultured podocytes in the presence of bone marrow conditioned media collected as described in materials and methods. Quantification of cells with intact stress fibers showed treatment of podocytes with BMC conditioned media causes actin depolarization and loss of stress fibers in podocytes (**Figure 8A**). When compared to podocytes cultured in presence of conditioned media from healthy BMCs, only 30% of podocytes treated with conditioned media from R-FSGS patients were positive for stress fibers (**Figure 8B**). Furthermore, we measured the levels of 40 different known inflammatory modulators in the conditioned media using a previously described platform for multiplex ELISA^30^ (MSD). We detected a significantly lower IL-12p70 in conditioned media from FSGS patients as compared to that from the controls. This is in accordance with the RNA sequencing results (**Figure 8C**). The levels of two chemokines were significantly different in cell culture supernatants of R-versus NR-FSGS BMNCs. MIP-1α was significantly upregulated and MDC was significantly lower in R-FSGS (Figure 8D and 8E).

## Discussion

FSGS is a glomerular disease characterized by proteinuria, frequent progression to end-stage-renal disease, and recurrence after kidney transplantation in ∼40% of patients which negatively impacts long-term allograft survival. Several mechanisms have been proposed to explain the recurrence of FSGS but haven’t translated into substantial advances in patient care and disease management. Genetic mutations in podocyte genes and circulating factors resulting from immune dysfunction represent some of the proposed mechanisms. In mice, bone marrow derived myeloid precursor cells were shown to be the source of pathogenic factor suPAR but the human equivalent of the proposed precursor cells does not exist. Hence, we profiled the bone marrow derived cells from R- and NR-FSGS patients along with control patients with end-stage renal disease due to other causes and healthy individuals by single cell RNA sequencing. Here we show that multiple cell types including neutrophils, T cells and B cells in bone marrow adopt an inflammatory phenotype in R-FSGS patients with an upregulation of cytokine signaling involving interleukins, interferons and TNF. Genes associated with elevated mitochondrial respiration were upregulated which can be associated with increased activity of T cells such as seen in SLE patients^36^. Indeed, T cells from R-FSGS patients’ bone marrow shows an upregulation of genes enriched in mitochondrial electron transport. Significant upregulation of *TNFRSF25* (TNF receptor superfamily member 25) was observed in R-FSGS MNCs. Knockout studies in mice have suggested the role of TNFRSF25 in the removal of self-reactive T cells in thymus by inducing apoptosis^37^ and treatment of cultured human podocytes with TNF-α caused a significant downregulation of *TNFRSF25* in microarray studies^38^.

An activation of apoptotic pathways was observed possibly mediated by the modulation of Jak/STAT or NFκB signaling both of which showed upregulation. We also identified an upregulation of genes involved in ribosome biogenesis in the mononuclear cells of the bone marrow of R-FSGS patients. Aberrant ribosome biogenesis has been shown to activate the p53-dependent apoptotic pathway^39^ which is also observed in R-FSGS cells. The mononuclear cells from bone marrow of R-FSGS patients show an upregulation of genes *BAG6, BCL3, DDIT4, PML* and *HIPK2* previously known to be involved in the p53 mediated apoptotic signaling^39^. Interestingly, disease-gene association analysis revealed that the upregulated genes closely correspond to the genes involved in Diamond-Blackfan anemia which is also a disease of aberrant ribosomal biogenesis^40^. Antigen presentation by MHC class I and II is upregulated in a small subset of neutrophils (subcluster 2) and it is enriched in cells from R-FSGS patients. We believe that is representative of the post-transplant state of the patients and its observed in enrichment of similar pathways in B cells also. Additionally, neutrophils display an upregulation of genes involved in glycolysis and gluconeogenesis. *AldoA* (aldolase A, fructose-bisphosphate) is a glycolytic flux monitor with additional function in controlling inflammasome activation via NLRP3 via AMPK-FOXO3 pathway and is upregulated in neutrophils originating from R-FSGS patients^41^. Dysregulated NLRP3 inflammasome activity results in uncontrolled inflammation in many chronic diseases. Some studies suggest that blocking glycolysis inhibits the activation of NLRP3 by inhibiting HK1 or PKM/PKM2^42^. Rituximab in addition to plasmapheresis or immunoadsorption has been shown to be effective in a subset of patients with recurrence of FSGS after transplant. However, neither the mechanism of efficacy nor the reason for non-response has been elucidated. We find activation of B cell receptor signaling and overall activation of adaptive immunity in a subset of R-FSGS patients with high serum suPAR levels. This may be a first glimpse into the heterogeneity of R-FSGS patients.

When we look at CD34+ progenitor cells, we find that they subcluster into all precursors for lymphoid, myeloid and erythroid cells and we see that only a small portion of the GMP population expresses PLAUR consistent with previous publications that have suggested that neutrophils are the primary source of suPAR.

To our knowledge this is the first study profiling the bone marrow transcriptional landscape in FSGS patients. Our study shows an altered transcriptional profile of bone marrow cell populations in R-FSGS patients. Additionally, the profiles are representative of an inflammatory phenotype concurrent with loss of B-cell population and neutrophils mimicking a situation where they are responding to cytokines. It has been recognized that virus associated FSGS can be caused by inflammatory cytokines and drug induced FSGS can occur due to medications such as interferon-*α* but here we show that in primary FSGS similar pathways are activated in the bone marrow MNC^43^. Conditioned media from *in vitro* culture of bone marrow mononuclear cells induced injury in podocytes providing evidence for the secretion of inflammatory modulators by MNCs in FSGS. MNCs from R-FSGS patients secrete higher levels of MIP-1α which is a chemotactic cytokine proposed as a biomarker for several inflammatory diseases^44^. Urinary excretion of MIP-1α reduced in chronic kidney disease patients given angiotensin receptor blocker irbesartan^45^. We also observe a significant reduction of IL-12p70 secretion from FSGS BMNCs. IL-12p70 is an immunomodulatory cytokine with cell type and context dependent function. In transplant, IL-12p70 induces CD4+CD25+ Tregs to promote tolerance^46, 47^. Mice deficient for the IL-12p70 receptor (*IL-12Rβ2*^*-/-*^) rapidly develop severe autoimmune disease suggesting that IL-12p70 reduction in FSGS is associated with dysregulated inflammation^48^. BMNCs from R-FSGS patients also secrete reduced levels of anti-inflammatory chemokine MDC/CCL22 canonically secreted by M2 macrophages. Furthermore, pseudotime analysis of each cell type shows a decision node at which the cells undergo transcriptional changes and result in two different trajectories associated with control/NR-FSGS cells or R-FSGS cells providing valuable insight into the gene expression changes leading to the recurrence of FSGS after kidney transplant.

## Data Availability

All data produced in the present study are available upon reasonable request to the authors

## Figure legends

**Figure 1.** Study design. Bone marrow aspirate from patients with ESRD due to biopsy proven primary FSGS (n=10) or controls with ESRD due to other non-FSGS causes were collected at UCSF. Aspirates from 5 age and gender matched healthy individuals were purchased from Lonza. Aspirate was centrifuged to collect total bone marrow cells or enriched for mononuclear cells by Ficoll-Paque density gradient. Equal number of cells from each individual were pooled to load 60,000 cells per lane for cDNA library preparation on 10X platform using chromium single cell 3’ v2 chemistry. Cells were subjected to single cell RNA sequencing on Illumina NovaSeq S2 using 150bp paired end sequencing with minimum read depth of 50,000 reads per cell. Each individual in the cohort was genotyped using Illumina Omniexpress SNP array kit. Computational tool “Freemuxlet” with SNPs included from 1000 genomes variant was used to cluster the sequences followed by mapping to their original source using the genotypes. Data was analyzed in python to generate UMAPs and perform differential gene expression and trajectory analysis. Bone marrow mononuclear cells were cultured *in vitro* and supernatant was collected for the validation of secreted inflammatory factors by ELISA and podocyte injury assay.

**Figure 2.** Single cell transcriptomes from bone marrow mononuclear cells and total bone marrow cells were analyzed by single cell RNA sequencing (scRNAseq). A) Unsupervised clustering of 35,229 mononuclear cells resulting in 26 distinct cell populations and annotated using canonical markers. B) UMAP displaying the distribution of 7 major cell types identified from (A). C) 15,376 single cell transcriptomes from all cells of bone marrow followed by unsupervised clustering resulted in 17 distinct cell populations. D) UMAP of 8 cell types including stromal cells identified based on canonical markers in (C). E and F) Top ten significantly expressed gene in each cluster of MNC and whole bone marrow cells has been shown to correlate with canonical markers of cell types assigned.

**Figure 3.** Transcriptomic differences between R-FSGS and NR-FSGS MNCs. A) Upregulated genes B) Downregulated genes C) Expression of FSGS associated genes D) Gene enrichment analysis using GO shows upregulation of pathways associated with B cells, T cells and neutrophils. E) Bone marrow aspirate and serum suPAR levels in R- FSGS patients separates the patients into two groups, with high and low suPAR respectively. F) Number of significantly altered transcripts in R-FSGS patients with high suPAR is approximately 4 times higher than that in patients with low suPAR. G) Dot plot representation of top ten significantly upregulated genes in R-FSGS patients with high suPAR. H) Trajectory analysis of neutrophils showing separation of high and low suPAR cells at branch point 1.

**Figure 4.** Unsupervised analysis of neutrophils shows downregulation of differentiation markers. A) Neutrophils in bone marrow resolve into 9 independent subclusters. B) Cluster 0 displays enrichment of bone marrow MNCs from R-FSGS patients. C) MNCs from R- FSGS patients in cluster 0 display upregulation of proteases CTSL, TIMP1 and MHC class II. D) Genes with highest fold increase in neutrophils from R-FSGS patients. E) Gene set enrichment with GO shows activation of neutrophils and inflammation associated phenotype.

**Figure 5.** Unsupervised analysis of T cells. A) UMAP shows 6 different populations of bone marrow T cells. B) Cluster 2 and Cluster 5 show enrichment of T cells from R-FSGS patients. C) Gene enrichment of differentially expressed genes in T cells from R-FSGS vs NR-FSGS patients using GO platform. D) Pseudotime trajectory using monocle 2 shows T cells from R-FSGS patients separate at branchpoint 1 from the control cells based on their transcriptional profiles. E) Heatmap showing top genes associated with the developmental changes in transcripts from R-FSGS and branch containing cells from control and NR-FSGS T cells.

**Figure 6.** Unsupervised analysis of B cells in bone marrow. A) Three populations of B cells expressing markers of Pre B, Pro B and plasma cells were distinguished. B) B cells from R-FSGS were found throughout the bone marrow MNC landscape. C) Upregulated genes in B cells from R-FSGS patients compared to that in NR-FSGS. D) Expression on genes upregulated in B cells from R-FSGS superimposed on the UMAP. E) Gene enrichment analysis reveals enrichment of several pathways in B cells of R-FSGS patients including upregulation of cytokine signaling. F) Pesudotime trajectory displays transcriptional changes in B cells leading to the phenotype observed in R-FSGS patients enriched in state 4. G) Heatmap showing transcriptionally altered genes at branch point 1.

**Figure 7.** Unsupervised analysis of CD34+ cells. A) CD34+ cells in bone marrow resolve into 4 independent subclusters. B) Cluster 1 displays enrichment of bone marrow MNCs from R-FSGS patients. C) Differential gene expression followed by functional enrichment of genes in cluster 1using Gene Ontology shows upregulated genes involved in antigen processing and presentation by MHC class I. D) Trajectory obtained from pseudotime analysis shows control cells differentiating from cells from R-FSGS patients at one branch point. E) Significant genes expressed before and after the branching at node 1 associated with the two different cell fates. LMPP: Lymphoid primed multipotent progenitor, MPP: hematopoietic stem cell/multipotent progenitor cell, MEP: megakaryocyte-erythrocyte progenitor cell.

**Figure S1**. Distribution of cells in each cluster from individual patients shows minimal batch effect. (A) Bar graph shows the distribution of bone marrow mononuclear cells from individuals in each group present in all clusters. (B) Bar graph showing the distribution of cells from each group in 17 clusters identified for whole bone marrow cells.

**Figure S2**. Soluble uPAR levels are higher in bone marrow aspirate and serum of ESRD patients. A) Bone marrow suPAR in healthy, NR-FSGS, R-FSGS, and control ESRD patients with renal failure due to non-FSGS causes. B) Bone marrow suPAR is slightly higher in R-FSGS patients but does not reach significance. C) Serum soluble uPAR is slightly higher in R-FSGS patients and shows variability among the individuals.

## References

1. Wen, Y., Shah, S. & Campbell, K.N. Molecular Mechanisms of Proteinuria in Focal Segmental Glomerulosclerosis. Front Med (Lausanne) 5, 98 (2018).

2. Kitiyakara, C., Eggers, P. & Kopp, J.B. Twenty-one-year trend in ESRD due to focal segmental glomerulosclerosis in the United States. Am J Kidney Dis 44, 815–825 (2004).

3. D’Agati, V.D., Kaskel, F.J. & Falk, R.J. Focal segmental glomerulosclerosis. N Engl J Med 365, 2398–2411 (2011).

4. Fogo, A.B. Causes and pathogenesis of focal segmental glomerulosclerosis. Nat Rev Nephrol 11, 76–87 (2015).

5. Francis, A., Trnka, P. & McTaggart, S.J. Long-Term Outcome of Kidney Transplantation in Recipients with Focal Segmental Glomerulosclerosis. Clin J Am Soc Nephrol 11, 2041–2046 (2016).

6. Kienzl-Wagner, K., Waldegger, S. & Schneeberger, S. Disease Recurrence-The Sword of Damocles in Kidney Transplantation for Primary Focal Segmental Glomerulosclerosis. Front Immunol 10, 1669 (2019).

7. Gallon, L., Leventhal, J., Skaro, A., Kanwar, Y. & Alvarado, A. Resolution of recurrent focal segmental glomerulosclerosis after retransplantation. N Engl J Med 366, 1648–1649 (2012).

8. Savin, V.J. et al. Circulating factor associated with increased glomerular permeability to albumin in recurrent focal segmental glomerulosclerosis. N Engl J Med 334, 878–883 (1996).

9. McCarthy, E.T., Sharma, M. & Savin, V.J. Circulating permeability factors in idiopathic nephrotic syndrome and focal segmental glomerulosclerosis. Clin J Am Soc Nephrol 5, 2115–2121 (2010).

10. De Vriese, A.S., Wetzels, J.F., Glassock, R.J., Sethi, S. & Fervenza, F.C. Therapeutic trials in adult FSGS: lessons learned and the road forward. Nat Rev Nephrol 17, 619–630 (2021).

11. Sellier-Leclerc, A.L. et al. A humanized mouse model of idiopathic nephrotic syndrome suggests a pathogenic role for immature cells. J Am Soc Nephrol 18, 2732–2739 (2007).

12. Hahm, E. et al. Bone marrow-derived immature myeloid cells are a main source of circulating suPAR contributing to proteinuric kidney disease. Nat Med 23, 100–106 (2017).

13. Zheng, G.X. et al. Massively parallel digital transcriptional profiling of single cells. Nat Commun 8, 14049 (2017).

14. Kang, H.M. et al. Multiplexed droplet single-cell RNA-sequencing using natural genetic variation. Nat Biotechnol 36, 89–94 (2018).

15. Wolock, S.L., Lopez, R. & Klein, A.M. Scrublet: Computational Identification of Cell Doublets in Single-Cell Transcriptomic Data. Cell Syst 8, 281–291 e289 (2019).

16. Osorio, D. & Cai, J.J. Systematic determination of the mitochondrial proportion in human and mice tissues for single-cell RNA-sequencing data quality control. Bioinformatics 37, 963–967 (2021).

17. Ding, J. et al. Systematic comparison of single-cell and single-nucleus RNA-sequencing methods. Nat Biotechnol 38, 737–746 (2020).

18. Zhang, B. et al. The lncRNA Malat1 is dispensable for mouse development but its transcription plays a cis-regulatory role in the adult. Cell Rep 2, 111–123 (2012).

19. Johnson, W.E., Li, C. & Rabinovic, A. Adjusting batch effects in microarray expression data using empirical Bayes methods. Biostatistics 8, 118–127 (2007).

20. Wolf, F.A., Angerer, P. & Theis, F.J. SCANPY: large-scale single-cell gene expression data analysis. Genome Biol 19, 15 (2018).

21. Hao, Y. et al. Integrated analysis of multimodal single-cell data. Cell 184, 3573–3587 e3529 (2021).

22. Satija, R., Farrell, J.A., Gennert, D., Schier, A.F. & Regev, A. Spatial reconstruction of single-cell gene expression data. Nat Biotechnol 33, 495–502 (2015).

23. Traag, V.A., Waltman, L. & van Eck, N.J. From Louvain to Leiden: guaranteeing well-connected communities. Sci Rep 9, 5233 (2019).

24. Hay, S.B., Ferchen, K., Chetal, K., Grimes, H.L. & Salomonis, N. The Human Cell Atlas bone marrow single-cell interactive web portal. Exp Hematol 68, 51–61 (2018).

25. Subramanian, A. et al. Gene set enrichment analysis: a knowledge-based approach for interpreting genome-wide expression profiles. Proc Natl Acad Sci U S A 102, 15545–15550 (2005).

26. Kuleshov, M.V. et al. Enrichr: a comprehensive gene set enrichment analysis web server 2016 update. Nucleic Acids Res 44, W90–97 (2016).

27. Trapnell, C. et al. The dynamics and regulators of cell fate decisions are revealed by pseudotemporal ordering of single cells. Nat Biotechnol 32, 381–386 (2014).

28. Qiu, X. et al. Reversed graph embedding resolves complex single-cell trajectories. Nat Methods 14, 979–982 (2017).

29. Cakir, B. et al. Comparison of visualization tools for single-cell RNAseq data. NAR Genom Bioinform 2, lqaa052 (2020).

30. Delville, M. et al. A circulating antibody panel for pretransplant prediction of FSGS recurrence after kidney transplantation. Sci Transl Med 6, 256ra136 (2014).

31. Mariani, L.H. et al. Precision nephrology identified tumor necrosis factor activation variability in minimal change disease and focal segmental glomerulosclerosis. Kidney Int 103, 565–579 (2023).

32. Korbet, S.M. Treatment of primary FSGS in adults. J Am Soc Nephrol 23, 1769–1776 (2012).

33. Alachkar, N. et al. Podocyte effacement closely links to suPAR levels at time of posttransplantation focal segmental glomerulosclerosis occurrence and improves with therapy. Transplantation 96, 649–656 (2013).

34. Gussen, H. et al. Correction to: Neutrophils are a main source of circulating suPAR predicting outcome in critical illness. J Intensive Care 7, 40 (2019).

35. Reggiani, F. & Ponticelli, C. Focal segmental glomerular sclerosis: do not overlook the role of immune response. J Nephrol 29, 525–534 (2016).

36. Doherty, E., Oaks, Z. & Perl, A. Increased mitochondrial electron transport chain activity at complex I is regulated by N-acetylcysteine in lymphocytes of patients with systemic lupus erythematosus. Antioxid Redox Signal 21, 56–65 (2014).

37. Wang, E.C. et al. DR3 regulates negative selection during thymocyte development. Mol Cell Biol 21, 3451–3461 (2001).

38. Rashmi, P. et al. Perturbations in podocyte transcriptome and biological pathways induced by FSGS associated circulating factors. Annals of Translational Medicine (in press).

39. Zhang, Y. & Lu, H. Signaling to p53: ribosomal proteins find their way. Cancer Cell 16, 369–377 (2009).

40. Iskander, D. et al. Single-cell profiling of human bone marrow progenitors reveals mechanisms of failing erythropoiesis in Diamond-Blackfan anemia. Sci Transl Med 13, eabf0113 (2021).

41. Bai, D. et al. ALDOA maintains NLRP3 inflammasome activation by controlling AMPK activation. Autophagy 18, 1673–1693 (2022).

42. Xie, M. et al. PKM2-dependent glycolysis promotes NLRP3 and AIM2 inflammasome activation. Nat Commun 7, 13280 (2016).

43. Cravedi, P., Kopp, J.B. & Remuzzi, G. Recent progress in the pathophysiology and treatment of FSGS recurrence. Am J Transplant 13, 266–274 (2013).

44. Sa, V.C. et al. The pattern of immune cell infiltration in chromoblastomycosis: involvement of macrophage inflammatory protein-1 alpha/CCL3 and fungi persistence. Rev Inst Med Trop Sao Paulo 49, 49–53 (2007).

45. Ni, J., Huang, H.Q., Lu, L.L., Zheng, M. & Liu, B.C. Influence of irbesartan on the urinary excretion of cytokines in patients with chronic kidney disease. Chin Med J (Engl) 125, 1147–1152 (2012).

46. Verma, N.D. et al. Interleukin-12 (IL-12p70) Promotes Induction of Highly Potent Th1-Like CD4(+)CD25(+) T Regulatory Cells That Inhibit Allograft Rejection in Unmodified Recipients. Front Immunol 5, 190 (2014).

47. Daniel, V., Sadeghi, M., Wang, H. & Opelz, G. CD4+CD25+Foxp3+IFN-gamma+ human induced T regulatory cells are induced by interferon-gamma and suppress alloresponses nonspecifically. Hum Immunol 72, 699–707 (2011).

48. Zhang, G.X. et al. Role of IL-12 receptor beta 1 in regulation of T cell response by APC in experimental autoimmune encephalomyelitis. J Immunol 171, 4485–4492 (2003).

